# Understanding COVID-19 admissions in the UK; Analysis of Freedom of Information Requests

**DOI:** 10.1101/2022.06.06.22276032

**Authors:** T. Jefferson, J Brassey, C Heneghan

## Abstract

**Background:** The progression and severity of the COVID 19 pandemic have been measured based on the daily and total numbers of cases, hospitalisations and deaths. We focused on the nature of hospitalisations from 2020 to 2022.

**Methods:** We analysed the role played by SARS-CoV-2 in the pandemic in the UK; we lodged FOI requests to Public Health Wales (PHW), Scotland (PHS), and Northern Ireland (PHA NI), the UK Health Security Agency (UKHSA) and NHS England. We asked for all-cause hospital admission monthly numbers reported by days of positivity to SARS-CoV-2 since admission from March 2020. We grouped replies by respondents. We considered any positive tests acquired from day 8 post admission as evidence of in hospital transmission.

**Results:** PHW, PHS and PHA NI, provided data within two months. The proportion of people admitted who became positive after eight or more days was 33% in Northern Ireland, 24% in Scotland and 45% in Wales. There are seasonal fluctuations reflecting community admissions but no evidence that the proportion of those infected in hospitals reduced over time. No authorities had viral load or symptoms data relating to their datasets. Given the limitations in PCR reporting, it is impossible to know how many “positive” cases were active. UKHSA did not hold the data, and NHS England did not clarify the content of its website.

**Conclusion:** Aggregate data of “cases of Covid” in hospitals should not be used to inform policy of decision-makers until coordination, and proper interpretation of the dataset are instigated.

## Introduction

Since the beginning of the Covid 19 pandemic, its progression and severity have been measured based on the daily and total numbers of cases, hospitalisations and deaths. We have already reviewed the science-based on case ascertainment, death attribution and excess deaths in nursing homes. We found that cases were undefined because of misuse and misreporting of tests. There are 14 different ways in which deaths were attributed. An unknown proportion of deaths in homes were due to neglect and not infections. [1]

The importance of hospital episodes to the Covid narrative is illustrated by the daily and continued briefings by the Chief Medical Officer and Chief Scientific Advisor stressing the importance of “hospital pressures”. [2-6]

Here we look at the role of hospital-acquired SARS-CoV-2 infection based on data either available or obtained from Freedom on Information (FOI) requests, ours or other people’s.

We, therefore, submitted FOI requests that asked: ‘Could you please provide me with hospital admission data reported by days of positivity to SARS-CoV-2 since admission back to March 2020. For clarity, if someone was positive when admitted, that would be Day 0, if found positive after 5 days, Day 5 and so on.’

## Methods

The FOIA 2000 allows for a public “right of access” to information held by public authorities in the UK. We used the website WhatDoTheyKnow to facilitate FOI requests to NHS England, the UK Health Security Agency and the Public Health Agency of Northern Ireland. The website forwards requests to the appropriate authority and publishes the subsequent responses. We used a similar approach in previous reports on the role of PCR testing and the attribution of deaths in the UK During the SARS-CoV-2 Pandemic. [7, 8]

We used the same email to make requests to Wales, which publish results on their government website through a Disclosure Log. [9] Requests were made by emailing. Similarly, in Scotland, after initial triage, requests were managed by the COVID-19 Data & Analytical Team of Public Health Scotland by emailing PHS.Covid19Data&.

We collated FOI responses to a preliminary understanding of the current knowledge of the role played by SARS-CoV-2 in hospital episodes and provide an interpretation of the various FOI responses and recommendations to improve the understanding of what is going on. First, a distinction must be made between those who were positive for Covid 19 on admission and those who subsequently tested positive. Given the incubation period of up to 7 days, we considered a conservative lag time of 7 days from the day of admission to testing positive as sufficient to distinguish between those who certainly or probably arrived in hospital after contact with the virus and those who were infected while in hospital.

The datasets we acquired were provided in Microsoft excel format. We used excel to summarise and describe the data and produce the graphs. Finally, we set out the results by each of the agencies, including detailed responses to the requests and analysis of the results where provided.

## Results

We received positive responses with data relating to our request from the Public agencies of Northern Ireland, Wales and Scotland. (See web Appendix 1 for the data) NHS England report they ‘do not hold granular hospital admission data reported by days of positivity to SARS-CoV-2 since admission.’ Similarly, the UKHSA reports ‘they do not hold the data.’

### NHS England

NHS England’s response reported they did not hold information for hospital admission data by days of positivity back to March 2020. (See box 1 Timeline of responses)

The response referred to NHS England’s COVID-19 Hospital Activity [10] which includes a monthly publication of COVID-19 data, a weekly publication of COVID-19 admissions and bed occupancy data and a daily publication of COVID-19 admissions and bed occupancy data. The data includes the following definitions [11]

- *Total reported admissions to hospital and diagnoses in hospital* (the number of patients admitted in the previous 24 hours where patients known to have COVID-19 plus patients diagnosed in hospital with COVID-19 in the previous 24 hours)
- *Estimated new hospital cases* (the number of patients admitted in the previous 24 hours for the first time with COVID-19 plus the number of patients diagnosed in hospital in the previous 24 hours)
- *Estimated new admissions to hospital from the community* (Shows the number of patients admitted in the previous 24 hours for the first time with COVID-19 plus the number of patients diagnosed in hospital in the previous 24 hours where the test was within 7 days of admission)
- *Estimated new hospital admissions from the community with 3-7 day lagging* (the number of patients admitted in the previous 24 hours for the first time with COVID-19 plus the number of patients diagnosed in hospital in the previous 24 hours where the test was within 48 hours of admission plus the number of patients diagnosed in hospital in the previous 24 hours where the test was 3-7 days after admission (lagged by 5 days)
- *Total reported hospital admissions and diagnoses from a care home* (the number of patients admitted in the previous 24 hours with COVID-19 or diagnosed with COVID-19 in the previous 24 hours where admitted from Care Homes)
- *Total beds - occupied by confirmed COVID-19 patients* (the number of beds containing confirmed COVID-19 patients

Also, information since June 2021 includes those primarily treated ‘for’ Covid and those ‘with’ Covid but for whom the primary reason for being in hospital was non-Covid related. Trusts are asked to provide “for” and “with” split on a ‘best endeavours’ basis.

We could not identify the data referred to in the FOI for being ‘diagnosed with COVID within 8 days after admission.’ (see Box 1 Timeline, May 12 response)

It is possible - although not straightforward - to estimate the probable number of hospital-acquired infections based on the number of new cases minus the estimated new admissions to hospitals from the community. In 2020, using this method, we reported that 18% of all new hospital cases on October 6, which in the North West of England, HCAIs made up 24% of all patients on this date. [12]

#### Box 1.

**Timeline of NHS England responses**

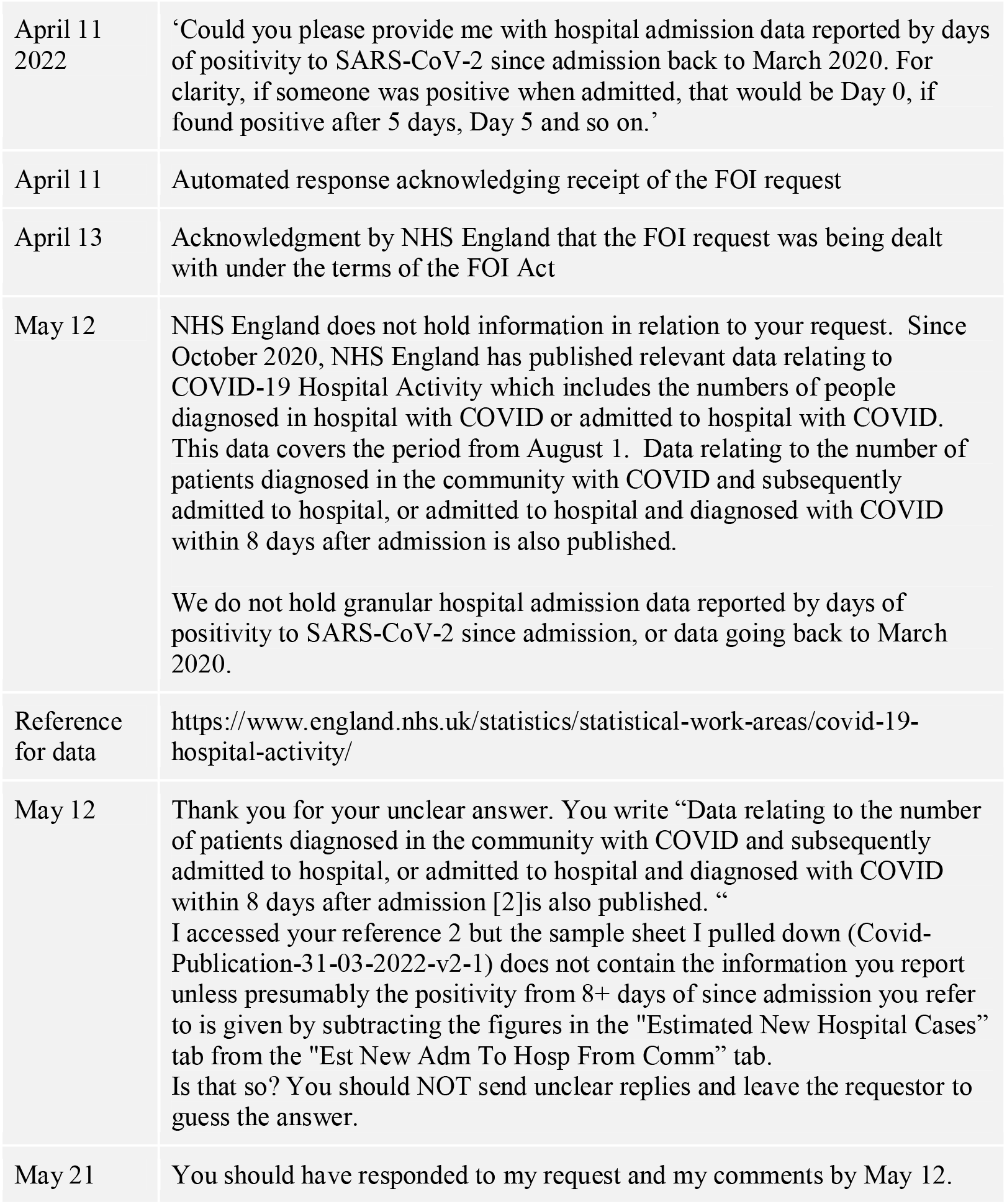

### UK Health Security Agency (UKHSA)

UKHSA’s response confirmed they do not hold the data.

The Office for National Statistics considers the UKHSA to hold the data on the cycle threshold (Ct) used on RT-PCR tests to determine a positive test in hospitals. [13] The UKHSA reports provide data on PCR positivity by date, reinfections, genomic sequencing, and cycle threshold analysis. It is unclear why they report they do not also hold the data by the day of the test result.

The UKHSA response was concerned about confidentiality. We consider this misplaced as their reports detail the ‘numbers of confirmed cases, outbreaks, hospitalisations and deaths - and where possible broken down by factors including age, gender, region and setting.’ [14] The claim of confidentiality is further undermined by UKHSA reports that are in the public domain of the median Ct value by the number of days since symptom onset in cases that have been sequenced (full sequencing or reflex assay). [15]

#### Box 2.

**Timeline of UKHSA responses [16]**

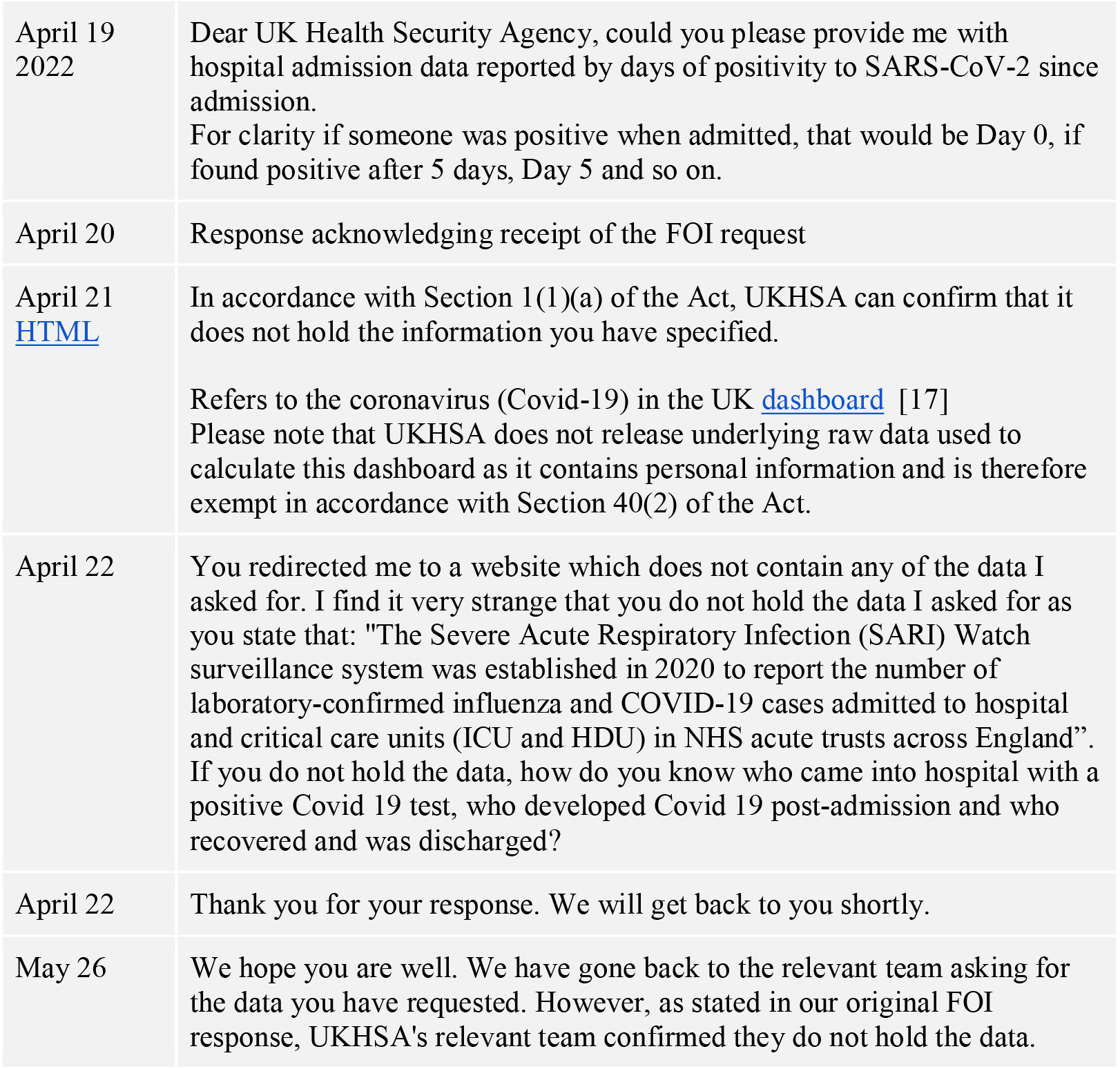

### Public Health Agency for Northern Ireland (PHA NI)

On May 23, the PHA NI responded positively to our request that included a downloadable excel file with the data for the positive test by day since admission. The results showed 21,813 first positive tests reported between 26 February 2020 and 18 May 2022. Of these, 10,174 (47%) patients had a positive test before admission, and 11,389 patients had a positive in hospital. Of these, 5,176 (51%) positive tests were on the day of admission, and one third were positive on day 8 or more (n=3,753, 33%).

#### Box 3.

**Timeline of Northern Ireland responses [18]**

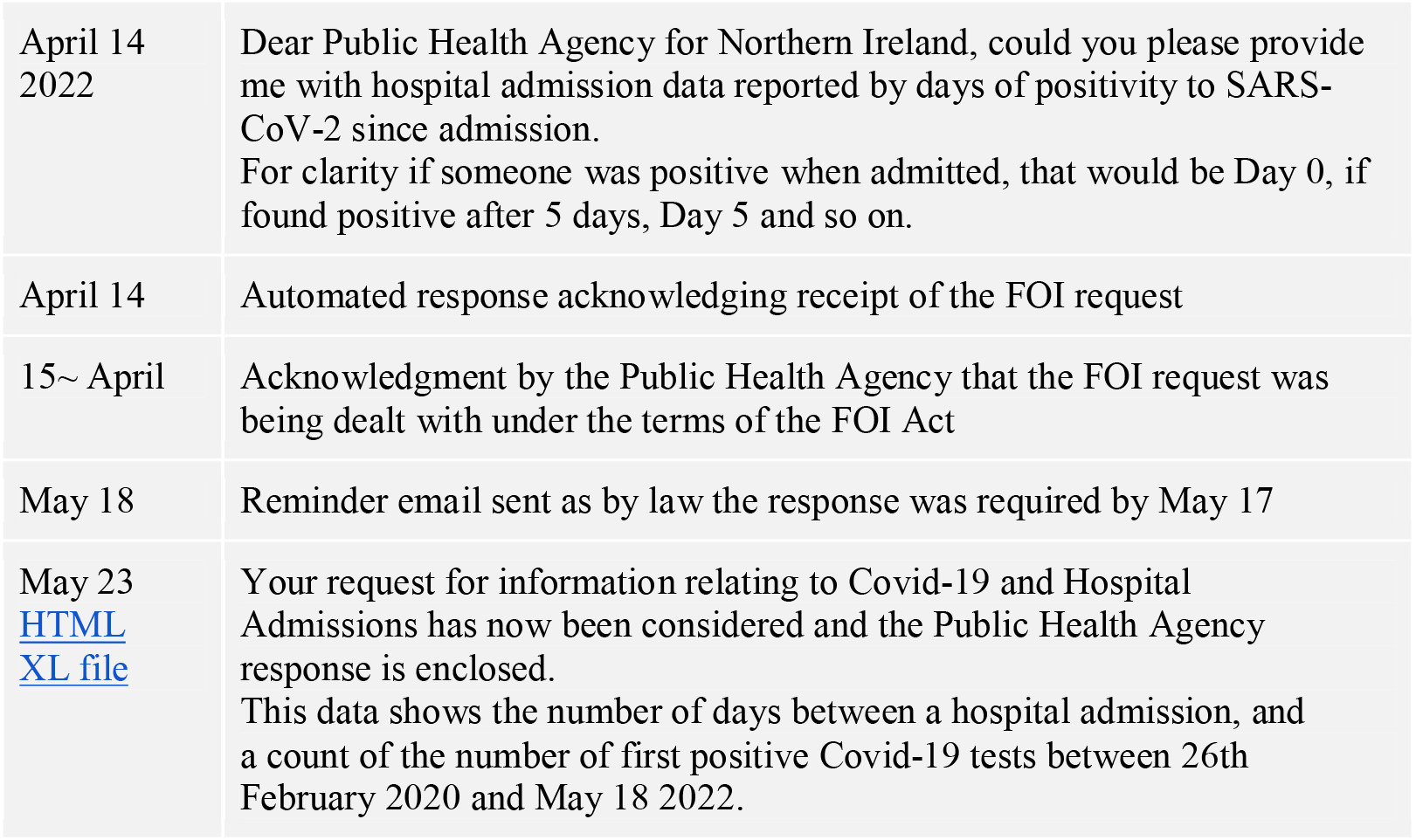

### Public Health Scotland (PHS)

PHS provided data from the period 1 March 2020 to 31 March 2022. The data included RAPID COVID-19 Admissions on individuals who first test positive for COVID-19 (including reinfections after 90 days) 14 days prior to or during their admission to a hospital. Only surgical, medical and paediatric specialities were included in the analysis.

RAPID data uses an estimated discharge date in cases where information is missing, using the following rules:

1) If an individual stay contains an episode submitted in the most recent extract, the date of discharge is calculated as the date of the latest admission occurring in the specific hospital plus one day;
2) If the stay does not contain an episode submitted in the most recent extract, then the date of discharge is computed as 4 days after the admission date for the last episode in the stay. If the last episode admission date is not available, the stay admission is used;
3) RAPID stays are single hospital stays and therefore may contain duplicates in cases where individuals move between hospitals.

### PHS Results

Table 1 shows PHS Scotland reported 57,255 positive tests with data relating to the day of hospital admission between March 2020 and March 2022.

**Table 1:** Public Health Scotland admissions data by Month.

|  | Community Onset<br>(Days -1 to -14) | Non-Hospital Onset<br>(Days 0 to 1) | Indeterminate Hospital Onset<br>(Days 2 to 7) | Probable Hospital Onset<br>(Days 8 to 14) | Definite Hospital Onset<br>(Days 15+) |
| --- | --- | --- | --- | --- | --- |
| Mar-20 | 217 | 1160 | 140 | 123 | 265 |
| Apr-20 | 714 | 2043 | 190 | 125 | 113 |
| May-20 | 337 | 330 | 42 | 25 | 18 |
| Jun-20 | 44 | 54 | 15 | * | 13 |
| Jul-20 | 9 | 11 | * | * | 25 |
| Aug-20 | 29 | 25 | * | * | 63 |
| Sept-20 | 165 | 157 | 33 | 47 | 210 |
| Oct-20 | 1438 | 1006 | 205 | 230 | 331 |
| Nov-20 | 1357 | 810 | 196 | 212 | 358 |
| Dec-20 | 1252 | 866 | 269 | 297 | 464 |
| Jan-21 | 2799 | 1474 | 422 | 335 | 242 |
| Feb-21 | 1353 | 709 | 123 | 97 | 52 |
| Mar-21 | 589 | 327 | 42 | 32 | 13 |
| Apr-21 | 245 | 167 | 16 | * | 20 |
| May-21 | 254 | 155 | 16 | * | 33 |
| Jun-21 | 564 | 405 | 39 | 28 | 57 |
| Jul-21 | 1209 | 811 | 49 | 14 | 85 |
| Aug-21 | 963 | 720 | 100 | 48 | 172 |
| Sept-21 | 2312 | 1305 | 160 | 106 | 180 |
| Oct-21 | 1637 | 1139 | 130 | 70 | 212 |
| Nov-21 | 1300 | 824 | 85 | 47 | 240 |
| Dec-21 | 1132 | 1115 | 170 | 194 | 444 |
| Jan-22 | 1646 | 1756 | 264 | 176 | 419 |
| Feb-22 | 812 | 1415 | 370 | 297 | 557 |
| Mar-22 | 1446 | 2879 | 748 | 448 | 408 |
| **Total** | 23823 | 21663 | 3824 | 2951 | 4994 |
| Percentage of total(n=57,255) | 41.6% | 37.8% | 6.7% | 5.2% | 8.7% |
| Percentage of in hospital total (n=33,432) |  | 64.8% | 11.4% | 8.8% | 14.9% |

\*Data suppressed due to low numbers

Of these, 23,823 (41.6%) were defined as community-onset (days -1 to -14).

A total of 33,432 tests were positive in the hospital setting. Of these, 21,663 (64.8%) were on days 0 to 1 (defined by PHS as non-hospital onset); 3,824 (11.4%) on days 2 to 7 (indeterminate); 2,951 (8.8%) on days 8 to 14 (probable) and 4,994 (14.9%) on days 15+ (definite hospital-onset).

Nearly one quarter (n=7,945, 23.8%) of hospital tests were positive after day 8 of admission. This percentage varied from 8.8% on April 20 to 9.8% in March 2021 to a high of 36.5% in Sep 2020, where full figures were available and not suppressed (See Figure 1). July 2020 through to December 2020 saw a high proportion of positive tests beyond day 8, with 2,237 (27.8%) positives reported out of 8,052 positives in hospital. The proportions fell to 9.4% in July 2021 and then rose in December 2021 to 24.9% (638/2561).

**Figure 1.**
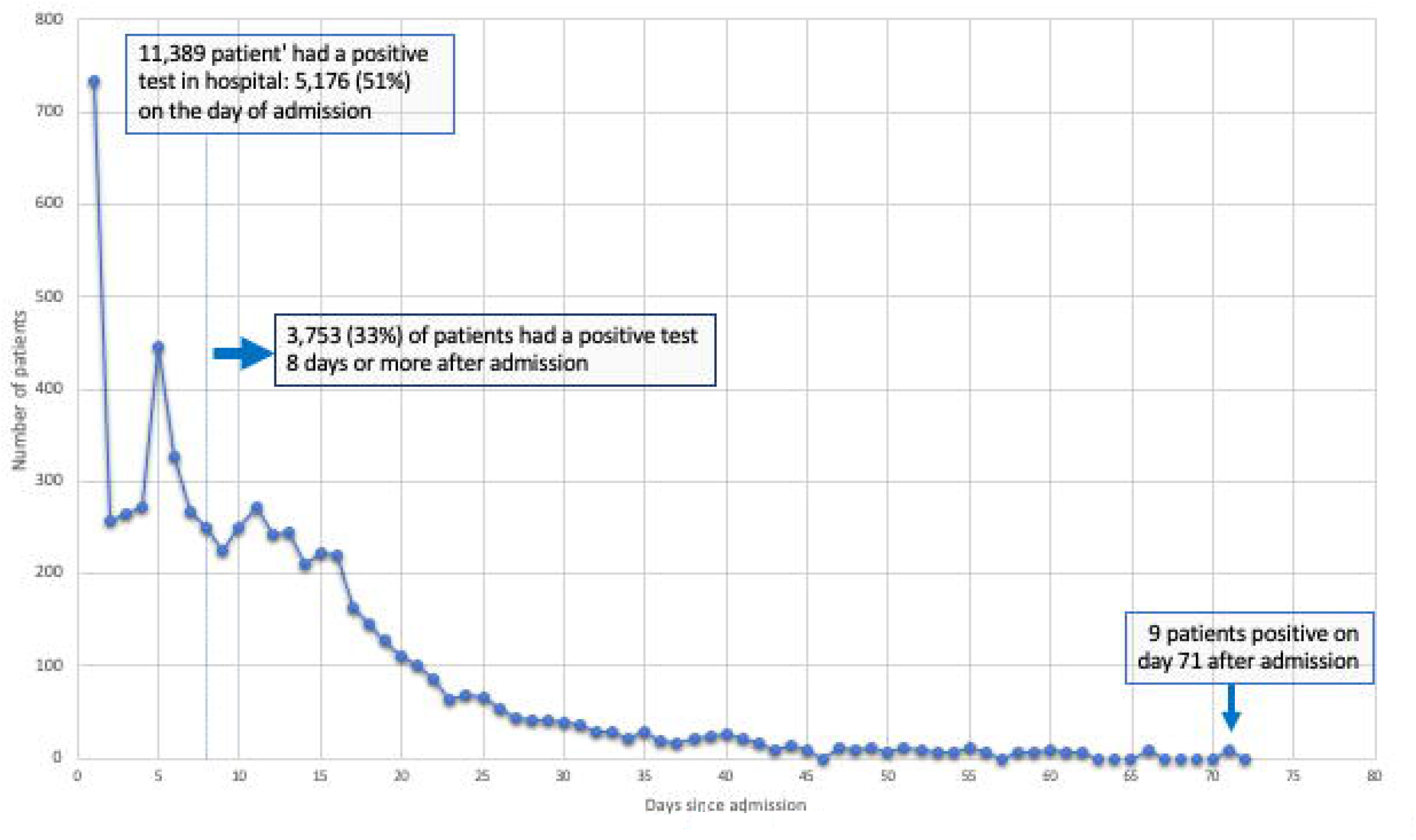
Public Health Agency for Northern Ireland Covid-19 and Hospital Admissions by day since admission 26 February 2020 to 18 May 2022.

#### Box 4.

**Timeline of Public Health Scotland responses**

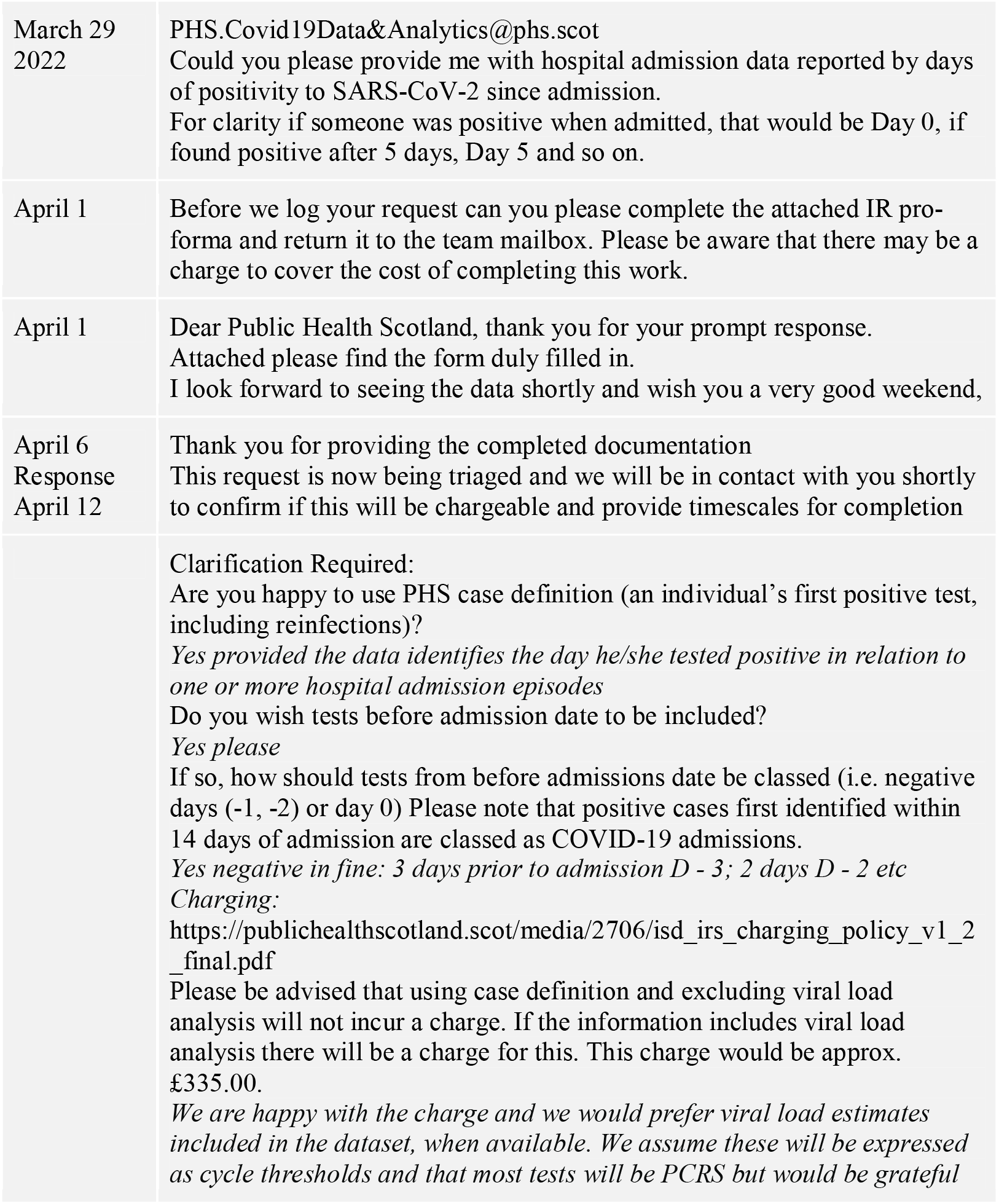
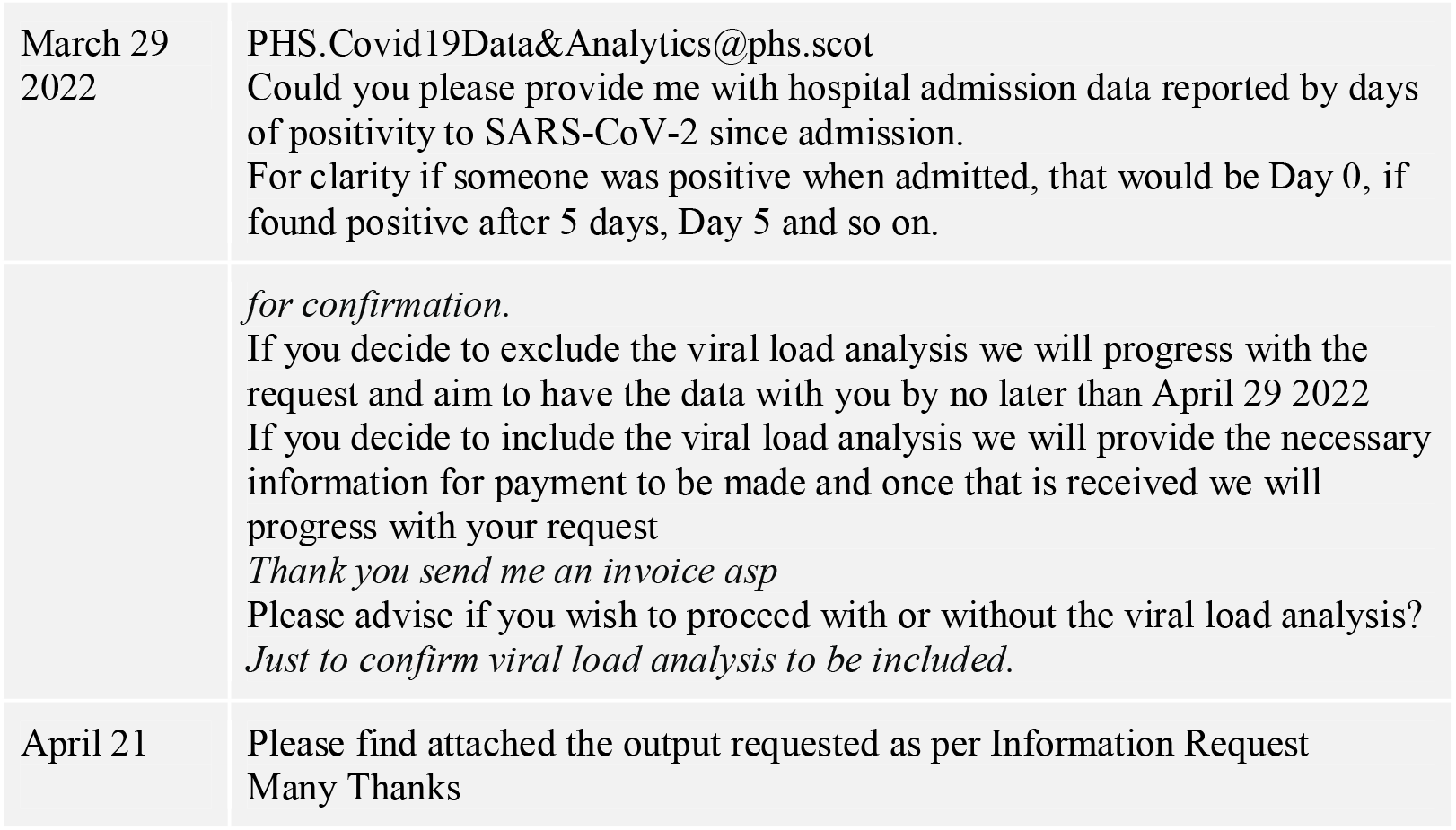

PH Scotland confirmed they did not hold viral load data in relation to the dataset released.

### Public Health Agency (PHW) Wales

PHW COVID-19 provided data on an individual with a PCR positive SARS-CoV-2 test where the sample was taken in a hospital in Wales between 1 March 2020 and 31 March 2022.

We assumed that the cases identified in the hospital were the first episode; however, it is unclear if the identified cases were readmissions of a previous episode. Hospital transfers within the same health board were classified as one continuous admission

PHW also provided weekly updated results on a tableau dashboard, [19] which stopped updating results on May 26 2022.

### PHW Results

Table 2 shows PHW reported 25,263 positive tests with data relating to the day of hospital admission between March 2020 and March 31 2022.

**Table 2.**
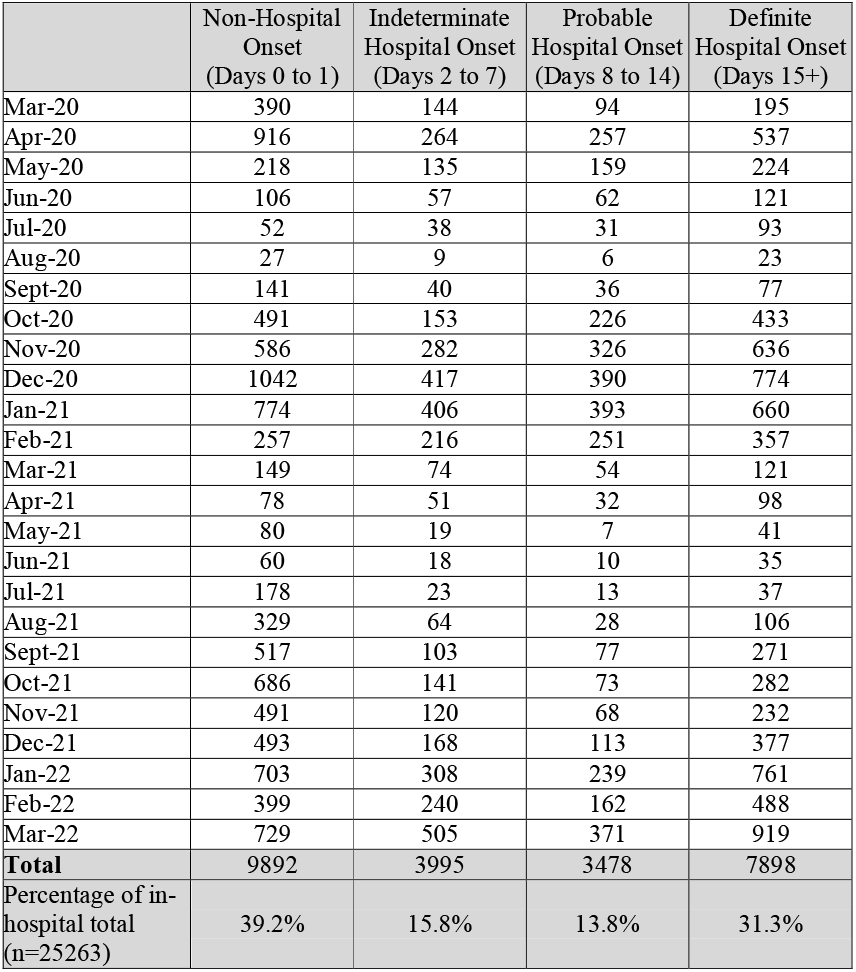
Public Health Wales admission data by Month.

Of the 25,263 tests positive in hospital, 9,892 (39.2%) were on days 0 to 1; 3,995 (15.8%) on days 2 to 7; 3,478 (13.8%) on days 8 to 14 and 7,898 (31.3%) on days 15+ [4,483 (17.7%) tests were reported as positive beyond 30 days of admission]

Therefore, approaching half (n=11,376, 45.1%) of the tests in hospital were positive after day 8 of admission. This percentage varied from 19.9 % in July to a high of 57.9% in July 2020 and 56.2% in February 2021. In March 2022, the number of positives after eight days of admission was above half (1290/2524, 51.1%) of the positive results (See Figure 2).

**Figure 2.**
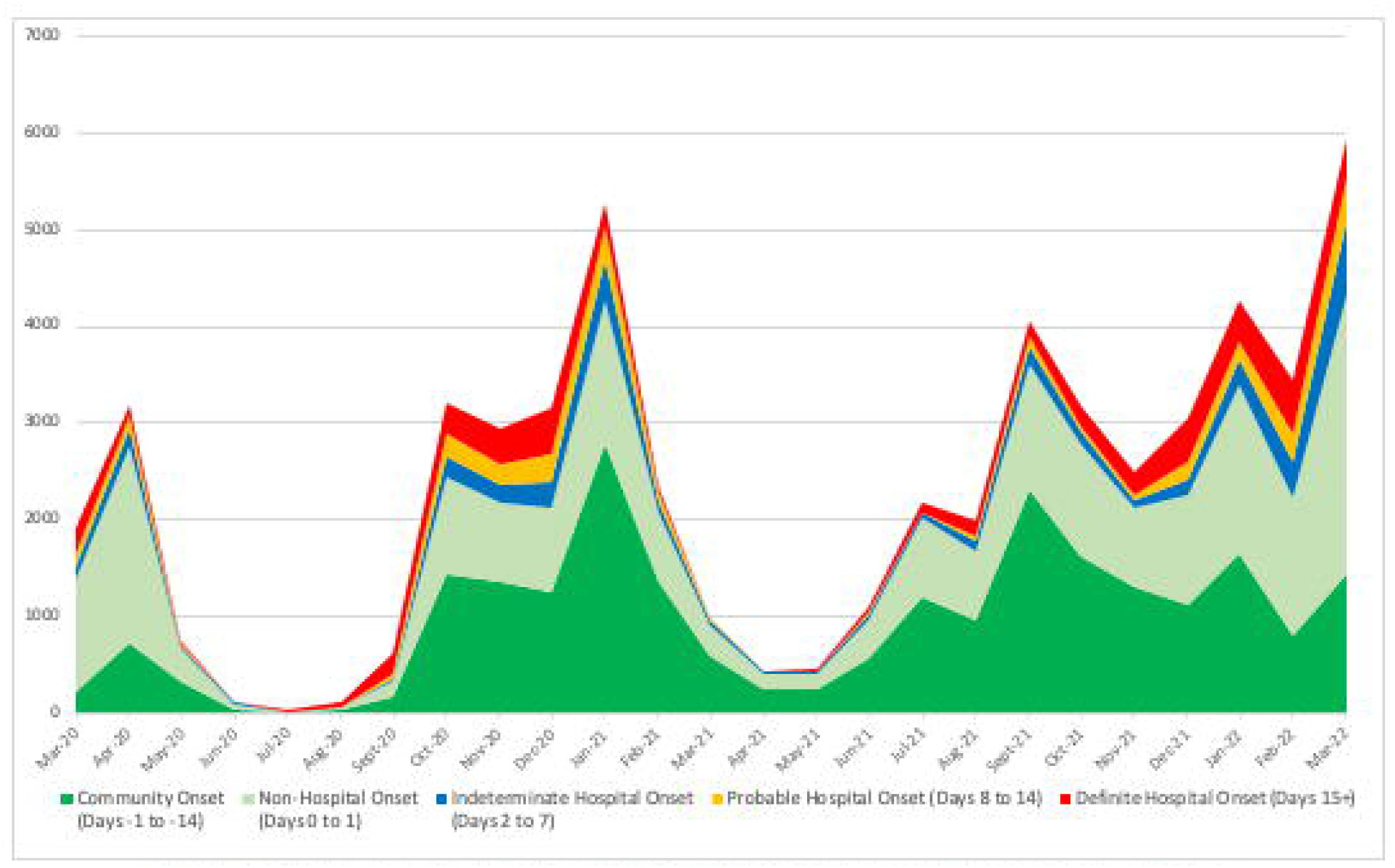
Public Health Scotland PCR positive tests by day of hospital admission March 2020 to 2022

**Figure 3.**
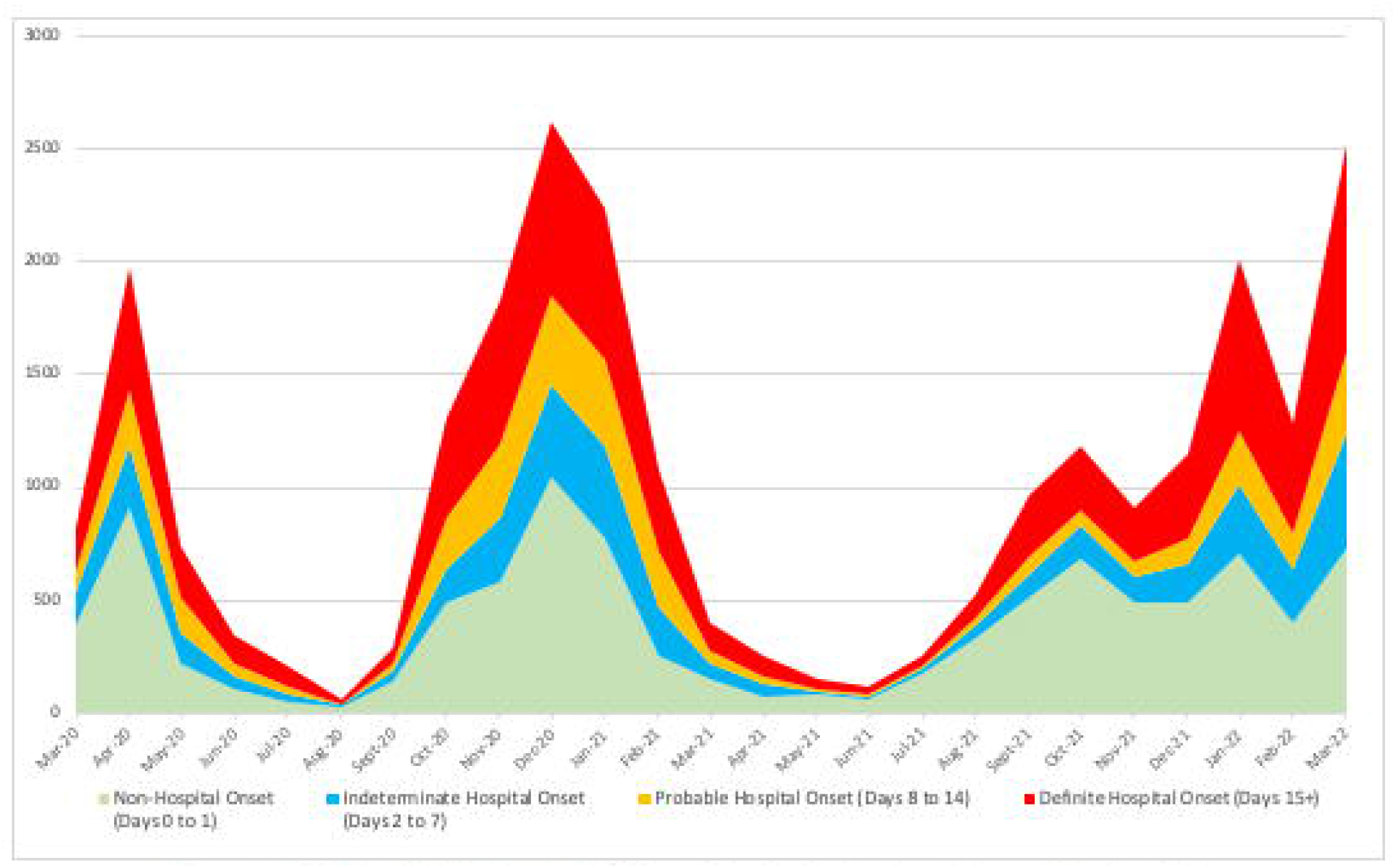
Public Health Wales PCR positive tests by day of hospital admission 1 March 2020 to 31March 2022

#### Box 5.

**Timeline of Public Wales responses**

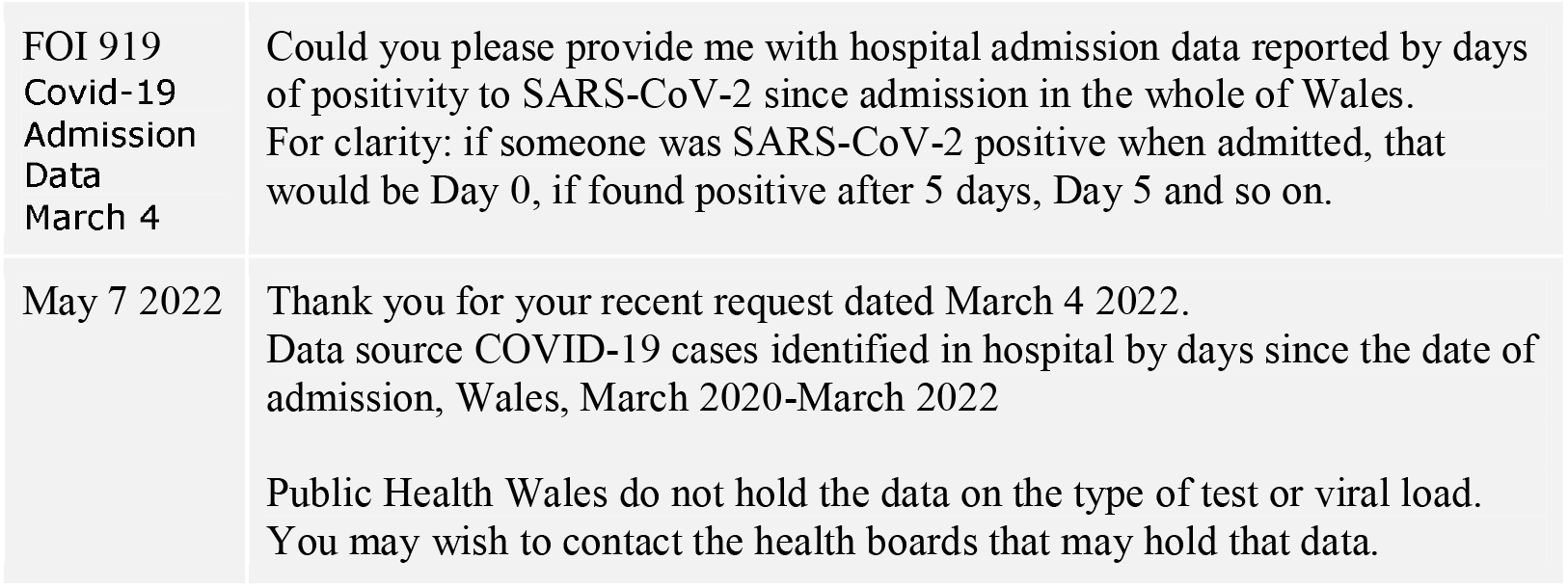

## Discussion

The “hospital pressures” concept is based on an aggregate total of cases admitted to hospital in a defined time frame. [20] Underpinning the pressure has been the daily and cumulative numbers of COVID-19 patients based on the assumption that all hospital PCR positive cases are admissions. [21]

However, a variable but high proportion of “new admissions” [22] in the devolved nations were infected in hospitals. These variations show a clear correlation with community viral circulation and seasonality, as seen in the figures and bear little relationship with the imposition of restrictions.

Our analysis shows that there was considerable variation in the proportion of infections acquired in hospitals where we received responses. In Northern Ireland, one third were PCR positive from day 8 onwards, In Scotland, the figure was roughly a quarter (24%), and in Wales, the proportion was approaching half (45%). Variation also occurred over the year, and there was no evidence that the proportion of those infected in hospitals reduced over time.

One devolved nation (Wales) routinely published data on COVID 19 Hospital Acquired Infections. [18] Although the Welsh Tableau reports a mixture of the incident and prevalent cases and requires considerable work to interpret, PHW were aware of the problem. If ever the Tableau is updated again, some effort needs to be devoted to enhancing its clarity. PHW deserves praise for their efforts, although it is unclear what action is being taken to reduce the burden of HAIs, which could be as high as over 50%.

Scotland and Northern Ireland provided the data we requested, albeit after assigning analysts to the task. The implication is that both bodies do not routinely monitor the burden of COVID 19 Hospital Acquired Infections. This is problematic given the high burden of hospitalised people infected post-admission. Nevertheless, Scotland’s analysis was a helpful improvement as it categorised the data into five categories providing additional data on community-onset (days -1 to -14) and methods and data by clinical speciality and by Lateral Flow Device or by PCR.

However, the response by UKHSA is at odds with that of the devolved nations and is contradicted by the routine reports that it generates.

It is not clear how UKHSA can make statements on COVID 19 Hospital Acquired Infections, which it does every week, if they do not have access to the data on PCR positivity in relation to the timing of admission. In addition, the UKHSA’s remit appears not to include the UK, as it does not include the three devolved nations or at least ignores their data holdings.

Redirecting enquiries to websites that do not hold the data but require complicated calculations is not helpful. Furthermore, the data collected on the NHS England’s website is classified as “management information”. It is collected daily with seemingly no checks and “no revisions” made to the data. Users of this data are warned that “any analysis of the data should be undertaken with this in mind.” [23] NHS England stated they publish aggregate up to 8 days post-admission and “do not hold granular hospital admission data reported by days of positivity to SARS-CoV-2 since admission, or data going back to March 2020”. The Accuracy of the data is questionable based on their responses and statements on the website. We consider the determination of the 8-day post-admission positivity to be beyond the reach of the public; therefore, redirections that require complicated assumptions and calculations do not meet the requirements of the FOI Act.

Limitations of this review are that none of the respondents could link positivity to measures of viral burden or COVID symptoms, so the concept of an active case will remain unproven. The lack of details on the test circumstances means we cannot rule out contamination as a cause of a proportion of positive cases. Our previous work on PCR testing (CG report 7) [24] shows that very few trusts/HAs hold viral load data, and none connect it in a systematic way to individuals, even if it’s just by age group and reason for admission.

Public Health bodies may lack the resources to link the likelihood of active disease to aggregate data and interpret their role as population-wide, not individual. This may be true, but a confident interpretation of “positive” as synonymous with “active” cannot be made if you are not certain that “positive” means “active” and not “contaminated”.

In addition, the data set may contain an aggregate of new and repeated admissions. Although we cannot rule this out according to the sources, the consistent trend, which reflects community viral circulation, makes double-counting seem less important.

Those who stay in hospital longer are at risk of infection, probably because they are sufferers of chronic and debilitating pathologies with weaker immune systems. The outcome of COVID 19 Hospital Acquired Infections is outside of the scope of this review but, given its importance, will be the subject of further investigation.

We recommend assessing whether structural or other factors are at play.

We also recommend the publication of hospital-acquired infections data by Trust to derive hypotheses on what is affecting the wide variations between devolved nations, whether the records are in part artefactual or variables like types of buildings, layout or practices may be at play. [25]

The use of aggregate data of “cases of Covid” in hospitals should not be used to inform policy of decision-makers until coordination, and proper interpretation of the dataset are instigated. Without it, the British public and politicians will continue to be misled.

## Conclusion

Freedom of Information requests provide a valuable source of information; however, data on the relationship of the day of admission to PCR Positivity should be routinely available, especially if the information is used to guide societal restrictions. The data from Northern Ireland, Wales and Scotland suggests somewhere between 25 and 50% of patients test positive on day 8 of admission or thereafter. Such variation and high rates of HAIs suggest there is a need for further investigations to determine what works in what settings to reduce the burden of infection, particularly for the most vulnerable.

The discrepancy in data availability between England and the devolved nations is concerning.

## Data Availability

All data analysed in the present study are available upon reasonable request to the authors or are already in the public domain

## Acknowledgements

We thank the Public Health of Northern Ireland, Public Health Wales, and Public Health Scotland for providing the data we requested.

The study was partly funded by Collateral Global, and CH received funding support from the NIHR School of Primary Care Research [Project 569].

## Competing Interests

TJ’s competing interests are accessible at: https://restoringtrials.org/competing-interests-tom-jefferson

CH holds grant funding from the NIHR School of Primary Care Research, the NIHR BRC Oxford, and the World Health Organization for a series of Living rapid reviews on the modes of transmission of SARs-CoV-2 reference WHO registration No2020/1077093. He has received financial remuneration from an asbestos case and given legal advice on mesh and hormone pregnancy test cases. He has received expenses and fees for his media work including occasional payments from BBC Radio 4 Inside Health and The Spectator. He receives expenses for teaching EBM and is also paid for his GP work in NHS out of hours. He has also received income from the publication of a series of toolkit books for appraising treatment recommendations in non-NHS settings and for undertaking evidence reviews. He is an advisor to Collateral Global, the Sir James Mackenzie Institute for Early Diagnosis at St Andrew’s University, the WHO’s International Clinical Trials Registry Platform (ICTRP) and is a member of the Board of Preventing Overdiagnosis.

JB is a major shareholder in the Trip Database search engine (www.tripdatabase.com) as well as being an employee. Trip has worked with a large number of organisations over the years, none have any links with this study. The main current projects are with AXA and SARS-CoV-2 (WHO Registration 2020/1077093-0) and is part of the review group carrying out rapid reviews for Collateral Global. He worked on Living rapid literature review on the modes of transmission of SARS-CoV-2 and a scoping review of systematic reviews and meta-analyses of interventions designed to improve vaccination uptake (WHO Registration 2021/1138353-0).

## Notes

### Competing Interest Statement

TJ competing interests are accessible at: https://restoringtrials.org/competing-interests-tom-jefferson
CH holds grant funding from the NIHR School of Primary Care Research, the NIHR BRC Oxford, and the World Health Organization for a series of Living rapid reviews on the modes of transmission of SARs-CoV-2 reference WHO registration No2020/1077093. He has received financial remuneration from an asbestos case and given legal advice on mesh and hormone pregnancy test cases. He has received expenses and fees for his media work including occasional payments from BBC Radio 4 Inside Health and The Spectator. He receives expenses for teaching EBM and is also paid for his GP work in NHS out of hours. He has also received income from the publication of a series of toolkit books for appraising treatment recommendations in non-NHS settings and for undertaking evidence reviews. He is an advisor to Collateral Global, the Sir James Mackenzie Institute for Early Diagnosis at St Andrew's University, the WHO's International Clinical Trials Registry Platform (ICTRP) and is a member of the Board of Preventing Overdiagnosis.
JB is a major shareholder in the Trip Database search engine (www.tripdatabase.com) as well as being an employee. Trip has worked with a large number of organisations over the years, none have any links with this study. The main current projects are with AXA and SARS-CoV-2 (WHO Registration 2020/1077093-0) and is part of the review group carrying out rapid reviews for Collateral Global. He worked on Living rapid literature review on the modes of transmission of SARS-CoV-2 and a scoping review of systematic reviews and meta-analyses of interventions designed to improve vaccination uptake (WHO Registration 2021/1138353-0).

